# Early and long-term risk of new-onset atrial fibrillation after transient ischemic attack

**DOI:** 10.1101/2021.03.05.21253024

**Authors:** Francisco Purroy, Mikel Vicente-Pascual, Gloria Arque, Mariona Baraldes-Rovira, Robert Begue, Joan Farre, Yhovany Gallego, M. Pilar Gil-Villar, Gerard Mauri, Nuria Montalà, Miriam Paul-Arias, Cristina Pereira, Coral Torres-Querol, Daniel Vazquez-Justes

**Author notes:** **Corresponding author:** Francisco Purroy, MD, PhD. Stroke Unit. Department of Neurology. Professor of the Universitat de Lleida. IRBLleida. Hospital Universitari Arnaude Vilanova de Lleida. Avda Rovira Roure, 80. Lleida 25198, Spain. /Fax 973248754. The authors declare no conflicts of interest.

## Abstract

**Background:** Transient ischemic attack (TIA) provides a unique opportunity to optimize secondary preventive treatments to avoid subsequent ischemic stroke (IS). Although atrial fibrillation (AF) is the leading cause of cardioembolism in IS and anticoagulation prevents stroke recurrence (SR), limited data exists about the risk of new-onset AF after TIA.

**Methods:** We carried out a prospective cohort study of consecutive patients with TIA from January 2006 to June 2010. The risk of new diagnosis of AF and SR was defined after a median follow-up time of 6.5 (5.0-9.6) years. In a subgroup of consecutive patients, a panel of biomarkers was assessed during the first 24 h of the onset of symptoms.

**Results:** 723 TIA patients were included. New-onset AF was diagnosed in 116 (16.0%) patients, 42 (36.2%) of them during admission. 204 (28.2%) patients were included in the biomarker substudy. New-onset AF was associated with sex (female) (hazard ratio [HR] 1.61 [95% CI, 1.07-2.41]), age (HR 1.05 [95% CI, 1.03-1.07]), previous ischemic heart disease (HR 1.84 [95% CI 1.15-2.97]) and cortical DWI pattern (HR 2.81 [95% CI 1.87-4.21]). In the Kaplan-Meier analysis, NT-proBNP ≥218.2 pg/ml (log-rank test P<0.001) was associated with significant risk of new-onset AF during the first five years of follow-up. Patients with a new diagnosis of AF after admission and before five years of follow-up had the highest risk of SR (P=0.002).

**Conclusion:** The risk of new diagnosis of AF after TIA is clinically relevant. We identified clinical, neuroimaging and biomarker predictors of new-onset AF.

## INTRODUCTION

Transient ischemic attack (TIA) provides a unique opportunity to optimize secondary preventive treatments and avoid subsequent ischemic stroke (IS)^1^. Up to 20% of ISs are preceded by a TIA^2^. This proportion could be significantly decreased with appropriate early management^3^. Although atrial fibrillation is the leading cause of cardioembolism in IS patients in both sexes^4 5^, it is frequently unrecognized at the time of the index event^6^. Indiscriminate anticoagulation for embolic strokes of undetermined source has not proven effective in different clinical trials^7 8^. In contrast, anticoagulation prevents stroke recurrence (SR) after IS caused by AF^4^. Therefore, the identification of AF after IS and TIA is critical^9^. A history of previous IS or TIA is a major SR risk factor in patients with AF^10^. Limited data exists about the risk of new-onset AF after TIA^11^, as early management has mainly focused on the detection and treatment of intracranial or extracranial large artery atherosclerosis (LAA), which is the main predictor of SR^12-14^. The practice of ECG monitoring is less common for patients with TIA than for those with IS^15^, and when TIA patients are admitted to stroke units they are monitored for significantly shorter periods than patients with IS^16^. A recent meta-analysis which focused only on TIA patients evidenced a lower AF detection rate than for IS and TIA cohorts^11^.

The aim of our study was to determine the risk of new-onset AF in a cohort of TIA patients with long-term follow-up. We describe clinical characteristics, neuroimaging features and biomarker patterns related to AF occurrence in order to identify those patients who would benefit from long-term cardiac monitoring. We also include a study of the prognostic repercussions of diagnostic delay.

## METHODS

### Data availability statement

Requests for access to the data reported in this article will be considered by the corresponding author.

### Design and study population

The REGITELL registry (Registro de pacientes con ictus transitorio de Lleida [in Spanish]) methodology has been described in detail previously^17 18^. Consecutive TIA patients were included from January 2006 to June 2010 following the Strengthening the Reporting of Observational Studies in Epidemiology (STROBE) statement^19^. TIA was defined as a reversible episode of neurological deficit of ischemic origin that was fully resolved within 24 h.^20^ All patients underwent standard routine diagnostic work-up which included 12-channel electrocardiography (ECG), transcranial and supra-aortic ultrasound test, head computerized tomography and laboratory tests. Clinical characteristics, previous diagnosis of AF, ABCD^2^ score^21^ and CHA_2_DS_2_-VASc risk score were recorded^10^. Any potential, but non-definitive, TIA events were defined by the presence of isolated, atypical symptoms such as: unsteadiness; diplopia; dysarthria; partial sensory deficit; and unusual cortical vision^22^. All cases were reviewed by the senior neurologist (FP) and classified by atherosclerosis and small vessel disease phenotypes according to the ASCOD classification^23^. Patients in whom cardiopathy was suspected underwent extended cardiological examinations. Those patients without contraindications were evaluated by cranial MRI that included diffusion-weighted imaging (DWI) sequences within 7 days of symptom onset. Presence of DWI abnormalities and patterns of acute ischemic lesions were also recorded^17 24^.

### New-onset AF definition and ECG procedures

AF was defined as a period of >30 seconds duration of an absolute arrhythmia without detectable P-wave activity and irregular RR interval^4^. The way AF was detected was recorded: ECG at emergency department, continuous electrocardiographic monitoring (CEM) or Holter ECG on admission, and ECG after clinical symptoms during the follow-up. Holter ECG was performed within the first 48 h after admission. Holter data were analyzed by a blinded cardiologist. CEM (GE Healthcare, Chicago, IL, USA) was started immediately after admission and maintained for three days. It included a special software for AF detection. Whenever AF was suspected, an experienced neurologist (FP, DVJ, MV or GM) reviewed the CEM report for confirmation or dismissal. When a doubt arose, the ECG was reviewed by a cardiologist. Cases of suspected abnormal baseline ECG were reviewed by the same team.

Structured clinical visits were performed by a stroke physician (FP, MBR, YG, MPGV, GM, DVJ, MVP or MP) during the follow-up period to determine the risk of further vascular events. SR was defined by the appearance of new focal symptoms or signs associated with acute ischemic changes shown in neuroimaging scans (CT or MRI) made at 7 days, 3 months, one year, five years and 10 years. If a patient moved out of the local area or if travel to the hospital was impossible, the follow-up was conducted by phone. Recurrent events and new diagnoses of AF were also actively identified by an annual review of electronic medical records^18^.

### Biomarkers substudy

From January 2008 to June 2010, blood samples were obtained by standard venipuncture within the first 24 h after the onset of symptoms at the emergency department admission to test a panel of biomarkers^25^ that included high-sensitivity C-reactive protein (hs-CRP), interleukin-1-alpha (IL-1 α), IL-6, tumor necrosis factor-alpha (TNF-α), neuron-specific enolase (NSE), S100b, N-terminal pro-B type natriuretic peptide (NT-proBNP), copeptin, adiponectin and neopterin. Plasma, serum and buffy coat were obtained after centrifugation at 3,000 g at 4° C for 10 min, and aliquoted into cryovials for immediate storage at −80 ⍰C (Plataforma Biobancos PT17/0015/0027). Hs-CRP, IL-6, NSE, S100b and NT-proBNP levels were assessed in plasma samples (Hoffmann-La Roche, Basel, Switzerland) by an electrochemical chemiluminescence immunoassay using the COBAS 6000 e601 (Hoffmann-La Roche, Basel, Switzerland) at the Medical Laboratory of the HUAV. Other determinations were performed at the Clinical Neurosciences laboratory of the IRBLleida (Institut de Recerca Biomedica de Lleida). In detail, IL-1 α and TNF-α were quantified in serum using a solid-phase sandwich enzyme-linked immune sorbent assay (ELISA), commercially available in BIONOVA. The absorbance was read in a spectrophotometer using 450 nm wavelength. Copeptin was measured with a chemiluminescence sandwich immunoassay, determined with an immunoluminometric assay (Thermo Scientific B.R.A.H.M.S CT-proAVP LIA). Neopterin was quantified by competitive immunoassay using high-affinity monoclonal antibody (IBL America, MN, USA), and adiponectin was quantified with a competitive enzyme immunoassay ADIPOQ (Human) ELISA kit (Abnova).

### Statistical analysis

First, we compared the baseline characteristics, ASCOD classification, presence and distribution of acute lesions in DWI, ABCD^2^ score^21^, CHA_2_DS_2_-VASc^10^, outcomes and 2 2biomarker levels between non-AF, previous AF and new-onset AF patient groups. Second, we performed the same comparison, but excluding previous AF patients. Third, we compared previous AF and new-onset AF patients. Finally, we analyzed differences between new-onset AF groups that took into account time to diagnosis: less than one year, between one year and less than five years, and five years or more after the onset of symptoms. The quantitative variables were compared using either the student’s T-test or the Mann-Whitney U test. The qualitative variables were compared using the chi-squared test or Fisher’s exact test when the expected cell frequency was <5. Biomarkers were not normally distributed (P-P plot), and values were expressed as median (interquartile range). A Bonferroni correction (a multiple-comparison correction) was applied to all the significant associations to reduce the risk of finding false-positive associations. To calculate the sensitivity and specificity for biomarker cut-off values which allowed to discriminate TIA patients with new-onset AF from TIA patients without AF, a receiver operator characteristic (ROC) analysis was performed. The cumulative risks of new-onset AF after excluding patients with previous AF during follow-up were estimated using a Kaplan-Meier analysis. The results were censored at the time of the outcome event, patient death, or the end of the follow-up period. Risks were compared using the log-rank test. Data on patients with no information at 10 years were censored at the time of the last available follow-up. A Cox proportional hazards multivariable analysis was performed including clinical variables (model 1), clinical variables and neuroimaging features (model 2), clinical variables and biomarker data (model 3), and clinical variables, neuroimaging features and biomarker data (model 4) to identify predictors of SR. It was also adjusted for patient characteristics that significantly predicted outcomes in univariate logistic regression models. We compared the risk of SR in subgroups of patients categorized according to the main identified predictors using a Kaplan-Meier analysis and the log-rank test. All the tests were 2-sided. Missing data was included as a random effect when fitting the model for multivariable analyses. The statistical analysis of the data was carried out using the SPSS statistical package, version 24.0. (SPSS, Chicago, IL, USA) and Graphpad software version 6 (LLC).

### Standard protocol approvals, registrations, and patient consents

Written informed consent or assent from relatives was obtained for all the participants. The study was approved by our local Ethics Committee: the “Comité d’Etica i Investigació Clínica de l’Hospital Universitari Arnau de Vilanova de Lleida”^18^.

## RESULTS

723 TIA patients were included in the analysis after excluding 48 patients who were diagnosed as mimics. In 85 (11.8%) patients, AF was documented before the index event (Figure 1). New-onset AF was diagnosed in 116 (16.0%) patients: 31 (26.7%) of them after performing an ECG at the emergency room and 11 (9.5%) during admission, 18 (15.5%) after admission and during the first year of the follow-up, 29 (25.0%) between one year and five years and 27 (23.3%) beyond five years of follow-up. CEM was performed on 288 (47.4%) of the 607 patients with no previous diagnosis of AF or with no AF documented at the emergency room, and 139 (22.9%) underwent Holter ECG recording on admission. AF was identified in only 10 (3.5%) patients in CEM and no Holter ECG identified an AF event. 204 (28.2%) patients were included in the biomarker substudy. Compared to the patients not included in the substudy, a higher proportion had a previous stroke (28 [13.5%] vs. 38 [7.4%]; P=0.010) and events lasting from 10 to 60 minutes (84 [41.0%] vs. 164 [32.4%]; P=0.002) (Supplementary Table I).

**FIGURE 1.**
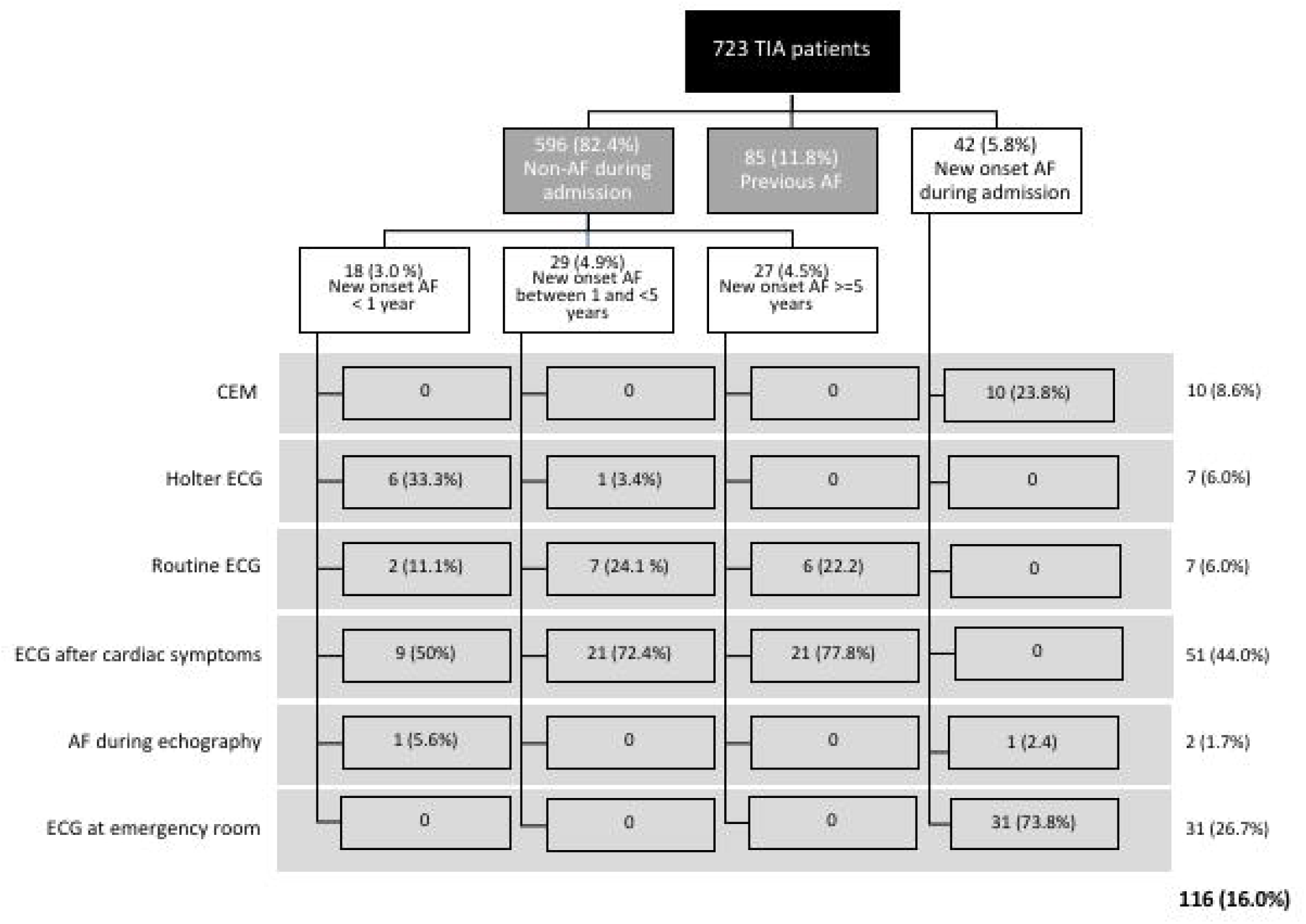
Proportion of patients with previous and new diagnosis of atrial fibrillation; and method of atrial fibrillation detection.

### Clinical and imaging characteristics of patients with new-onset atrial fibrillation

Like patients with previously diagnosed AF, patients with new-onset AF were older, had a higher proportion of carotid territory events, speech impairment and previous ischemic heart disease, but a lower proportion of non-definitive TIA events and isolated sensory impairment than patients without AF (Table 1). The proportion of women, positive DWI and cortical DWI patterns were higher in AF groups than non-AF patient groups. In addition, the CHA_2_DS_2_-VASc score was higher in AF groups than non-AF groups. No difference was observed in the risk of SR between groups. AF groups had significantly higher levels of NT-proBNP and adiponectin than non-AF groups.

**Table 1.** Clinical characteristics, neuroimaging features, outcomes and biomarker level by previous and new diagnosis of atrial fibrillation.

|  | All | No AF | Previous AF | New diagnosis of AF | p-value comparison between all groups | p-value comparison between no previous AF and new diagnosis of AF | p-value comparison between previous AF and new diagnosis of AF |
| --- | --- | --- | --- | --- | --- | --- | --- |
| Total, n (%) | 723 | 522 (72.2) | 85 (11.8) | 116 (16.0) | ... | ... | ... |
| Years of follow-up, median (IQR) | 6.5 (5.0-9.6) | 6.6 (5.1-9.8) | 5.9 (3.3-8.1) | 6.7 (5.1-9.9) | 0.003 | 0.458 | 0.020 |
| Sex female, n (%) | 302 (41.8) | 200 (38.3) | 38 (44.7) | 64 (55.2) | 0.003* | 0.001* | 0.095 |
| Age, mean (SD) years | 70.7 (11.9) | 68.9 (12.4) | 76.8 (8.0) | 74.6 (9.9) | <0.001* | <0.001* | 0.143 |
| Previous ischemic stroke n (%) | 66 (9.1) | 50 (9.6) | 8 (9.4) | 8 (6.9) | 0.660 | 0.363 | 0.515 |
| Hypertension | 481 (66.5) | 328 (62.8) | 70 (82.4) | 83 (71.6) | 0.001* | 0.076 | 0.076 |
| Alcoholism, n (%) | 21 (2.9) | 18 (3.4) | 1 (1.2) | 2 (1.7) | 0.364 | 0.335 | 1.000 |
| Previous ischemic heart disease, n (%) | 99 (13.7) | 54 (10.3) | 18 (21.2) | 27 (23.3) | <0.001* | <0.001* | 0.724 |
| Diabetes mellitus, n (%) | 215 (29.7) | 158 (30.3) | 26 (30.6) | 31 (26.7) | 0.739 | 0.450 | 0.548 |
| Active smoking, n (%) | 102 (14.1) | 86 (16.5) | 3 (3.5) | 13 (11.2) | 0.004 | 0.156 | 0.047 |
| Previous peripheral artery disease, n (%) | 26 (3.6) | 24 (4.6) | 0 (0) | 2 (1.7) | 0.054 | 0.157 | 0.224 |
| Hypercholesterolemia, n (%) | 246 (34.0) | 170 (32.6) | 31 (36.5) | 45 (38.8) | 0.388 | 0.199 | 0.737 |
| Previous heart failure, n (%) | 31 (4.3) | 12 (2.3) | 14 (16.5) | 5 (4.3) | <0.001* | 0.224 | 0.004 |
| Characteristics of the event |  |  |  |  |  |  |  |
| Duration of the event ≥ 60', n (%) | 377 (53.0) | 265 (51.9) | 49 (57.6) | 63 (54.8) | 0.065 | 0.525 | 0.300 |
| Multiple events, n (%) | 165 (22.8) | 128 (24.5) | 12 (14.1) | 25 (21.6) | 0.099 | 0.498 | 0.179 |
| Carotid territory event, n (%) | 367 (50.8) | 242 (46.4) | 55 (64.7) | 70 (60.3) | 0.001* | 0.006 | 0.529 |
| Vertebrobasilar event, n (%) | 78 (10.8) | 58 (11.1) | 11 (12.9) | 9 (7.8) | 0.456 | 0.287 | 0.225 |
| Undetermined territory event, n (%) | 286 (39.6) | 228 (43.7) | 21 (24.7) | 37 (31.9) | 0.001* | 0.020 | 0.266 |
| Possible, non-definitive TIA event, n (%) | 79 (10.9) | 72 (13.8) | 3 (3.5) | 4 (3.4) | <0.001* | 0.002* | 0.975 |
| Speech impairment, n (%) | 449 (62.1) | 302 (57.9) | 62 (72.9) | 85 (73.3) | 0.001* | 0.002* | 0.958 |
| Motor impairment, n (%) | 377 (52.1) | 271 (51.9) | 44 (51.8) | 62 (53.4) | 0.954 | 0.765 | 0.813 |
| Isolated sensory impairment, n (%) | 53 (7.3) | 47 (9.0) | 1 (1.2) | 5 (4.3) | 0.015 | 0.095 | 0.197 |
| Campimetric visual deficit, n (%) | 22 (3.0) | 12 (2.3) | 5 (5.9) | 5 (4.3) | 0.141 | 0.225 | 0.613 |
| ABCD<sup>2</sup> score, median (IQR) | 5.0 (4.0-6.0) | 5.0 (4.0-6.0) | 5.0 (4.0-6.0) | 5.0 (4.0-6.0) | 0.500 | 0.840 | 0.616 |
| Missing | 12 |  |  |  |  |  |  |
| CHA<sub>2</sub>DS<sub>2</sub>-VASC, mean (SD) | 3.0 (1.6) | 2.7 (1.7) | 3.7 (1.3) | 3.4 (1.6) | <0.001* | <0.001* | 0.258 |
| ASCOD classification, n (%) |  |  |  |  |  |  |  |
| A1 or 2 | 144 (19.9) | 115 (22.0) | 11 (12.9) | 18 (15.5) | 0.065 | 0.118 | 0.608 |
| A0 or 3 | 579 (80.1) | 407 (78.0) | 74 (87.1) | 98 (84.5) |  |  |  |
| S1 or 2 | 109 (15.1) | 100 (19.2) | 2 (2.4) | 7 (6.0) | <0.001* | 0.001* | 0.212 |
| S0 or 3 | 614 (84.9) | 422 (80.8) | 83 (97.6) | 109 (94.0) |  |  |  |
| Neuroimaging features |  |  |  |  |  |  |  |
| Positive DWI, n (%) | 244 (39.7) | 158 (34.9) | 28 (43.8) | 58 (59.8) | <0.001* | <0.001* | 0.046 |
| Missing | 109 |  |  |  |  |  |  |
| Pattern of DWI lesion, n (%) |  |  |  |  |  |  |  |
| Scattered in one vascular territory | 92 (37.6) | 53 (33.5) | 17 (60.7) | 22 (37.3) | 0.024 | 0.606 | 0.040 |
| Cortical lesion in one vascular territory | 73 (29.8) | 43 (27.2) | 6 (21.4) | 24 (40.7) | 0.082 | 0.056 | 0.078 |
| Multiple vascular territories | 19 (7.8) | 12 (7.6) | 2 (7.1) | 5 (8.5) | 0.969 | 0.830 | 0.831 |
| Subcortical | 61 (24.9) | 50 (31.6) | 3 (10.7) | 8 (13.6) | 0.004 | 0.007 | 0.709 |
| Follow-up events |  |  |  |  |  |  |  |
| Ischemic stroke recurrence, n (%) | 98 (13.6) | 65 (12.5) | 12 (14.1) | 21 (18.1) | 0.271 | 0.107 | 0.451 |
| Biomarker results; median (IQR) |  |  |  |  |  |  |  |
| NSE, pg/ml | 7.3 (5.0-10.0) | 6.9 (4.8-9.7) | 6.7 (4.9-11.0) | 7.5 (5.0-10.3) | 0.146 | 0.069 | 0.914 |
| Hs-CRP, mg/l | 3.6 (1.5-9.6) | 3.2 (1.4-9.3) | 5.2 (1.9-17.7) | 2.7 (1.3-13.0) | 0.228 | 0.970 | 0.182 |
| IL-6, pg/ml | 3.9 (1.8-8.5) | 3.4 (1.5-7.5) | 5.5 (1.9-10.1) | 4.6 (2.0-7.3) | 0.119 | 0.517 | 0.228 |
| NT-proBNP, pg/ml | 227.3 (76.5-561.8) | 161.3 (57.2-377.9) | 1416.0 (510.4-2507.5) | 466.4 (218.7-1031.0) | <0.001* | <0.001* | 0.009 |
| Neopterin, nmol/l | 13.4 (8.4-21.5) | 12.4 (8.1-17.4) | 8.6 (6.0-15.5) | 11.9 (9.6-20.2) | 0.493 | 0.550 | 0.254 |
| Copeptin, pmol/l | 9.8 (6.5-17.6) | 8.4 (5.6-15.1) | 11.7 (6.5-28.7) | 9.4 (5.8-10.5) | 0.260 | 0.288 | 0.405 |
| Adiponectin, microg/ml | 10.3 (7.1-14.3) | 8.6 (6.0-12.9) | 11.5 (8.5-18.3) | 11.7 (8.2-17.8) | 0.004 | 0.030 | 0.433 |
| TNF-α, pg/ml | 87.0 (63.5-144.5) | 81.0 (62.3-132.8) | 131.0 (55.0-147.5) | 93.0 (69.0-151.0) | 0.578 | 0.288 | 0.595 |
| IL-1 $\alpha$, pg/ml | 98.0 (68.0-161.0) | 90.5 (67.3-150.0) | 146.0 (64.5-166.0) | 109.0 (75.0-169.0) | 0.498 | 0.224 | 0.746 |
| S100b, pg/ml | 36.9 (26.5-52.7) | 34.5 (25.6-49.9) | 40.9 (23.8) | 31.8 (23.7-65.1) | 0.541 | 0.963 | 0.299 |

IQR, interquartile range; SD, standard deviation; DWI, diffusion-weighted imaging; ASCOD (A, atherosclerosis; S, small vessel disease; C, cardiac pathology; O, other causes; and D, dissection) classification. hs-CRP, high-sensitivity C-reactive protein; IL, interleukin; TNF-α , tumor necrosis factor-alpha; NSE, neuron-specific enolase; NT-proBNP, N-terminal pro-B type natriuretic peptide
\* Statistical significance after Bonferroni adjustment

When we compared previous and new-onset AF groups we detected a higher proportion of women in the new-onset AF group (38 [44.7%] vs. 64 [55.2%]; P=0.095), of previous heart failure (14 [16.5%] vs. 5 [4.3%]; P=0.004) and of positive DWI (28 [43.8%] versus 58 [59.8%]; P<0.001). Hypertension was more frequent in previous AF patients (70 [82.4%] vs. 83 [71.6%]; P=0.095). Finally, lesions on DWI classified as scattered in one vascular territory pattern were more frequent in the previous AF group than in the new-onset AF group (17 [60.7%] vs. 22 [37.3%]; P=0.040). In contrast, cortical lesion in one vascular territory was present in a greater proportion in new-onset AF patients (6 [21.4%] vs. 24 [40.7%]; P=0.078. Finally, previous AF patients had higher levels of NT-proBNP than new-onset AF patients (median [IQR]: 1416.0 [510.4-2507.5] vs. 466.4 [218.7-1031.0]; P=0.009).

When we analyzed the different categories of new-onset AF with respect to time of diagnosis from the onset of symptoms (Table 2), we identified a higher proportion of patients with cortical DWI pattern and S1 or S2 ASCOD grades in patients with new-onset AF detected ≥ 5 years after the onset of symptoms. In addition, these patients had lower levels of NT-proBNP and copeptin.

**Table 2.**
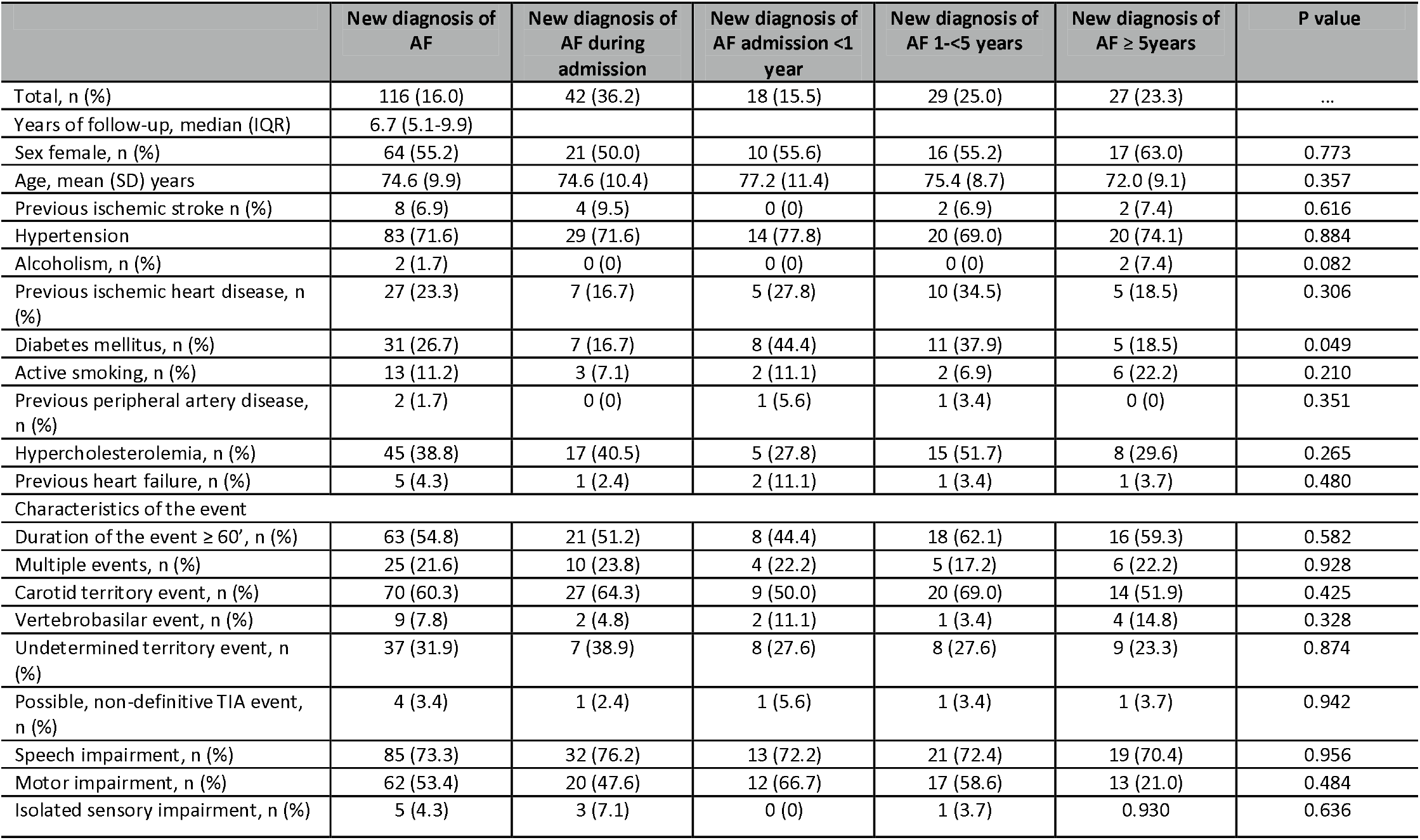
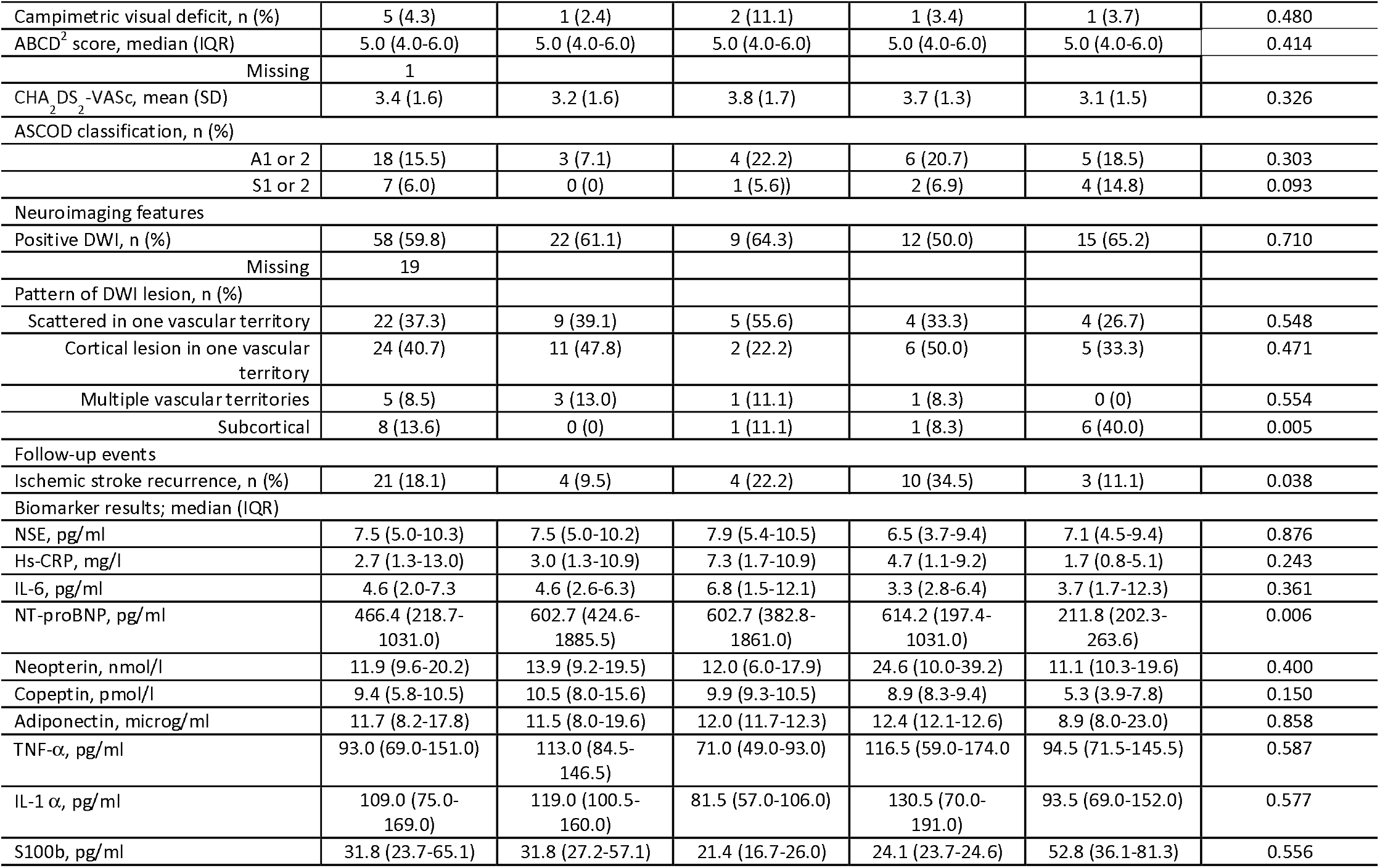
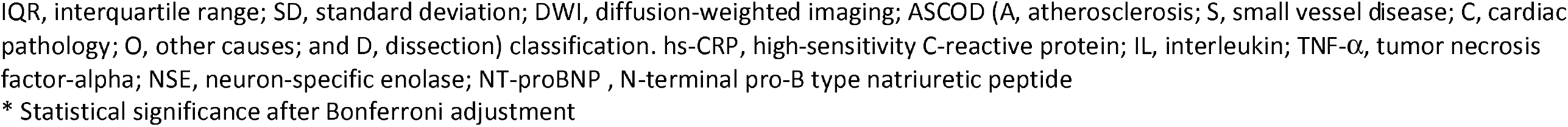
Clinical characteristics, neuroimaging features, outcomes and biomarker levels in new-onset atrial fibrillation groups by time to diagnosis.

|  | New diagnosis of AF | New diagnosis of AF during admission | New diagnosis of AF admission <1 year | New diagnosis of AF 1-<5 years | New diagnosis of AF ≥ 5years | P value |
| --- | --- | --- | --- | --- | --- | --- |
| Total, n (%) | 116 (16.0) | 42 (36.2) | 18 (15.5) | 29 (25.0) | 27 (23.3) | ... |
| Years of follow-up, median (IQR) | 6.7 (5.1-9.9) |  |  |  |  |  |
| Sex female, n (%) | 64 (55.2) | 21 (50.0) | 10 (55.6) | 16 (55.2) | 17 (63.0) | 0.773 |
| Age, mean (SD) years | 74.6 (9.9) | 74.6 (10.4) | 77.2 (11.4) | 75.4 (8.7) | 72.0 (9.1) | 0.357 |
| Previous ischemic stroke n (%) | 8 (6.9) | 4 (9.5) | 0 (0) | 2 (6.9) | 2 (7.4) | 0.616 |
| Hypertension | 83 (71.6) | 29 (71.6) | 14 (77.8) | 20 (69.0) | 20 (74.1) | 0.884 |
| Alcoholism, n (%) | 2 (1.7) | 0 (0) | 0 (0) | 0 (0) | 2 (7.4) | 0.082 |
| Previous ischemic heart disease, n (%) | 27 (23.3) | 7 (16.7) | 5 (27.8) | 10 (34.5) | 5 (18.5) | 0.306 |
| Diabetes mellitus, n (%) | 31 (26.7) | 7 (16.7) | 8 (44.4) | 11 (37.9) | 5 (18.5) | 0.049 |
| Active smoking, n (%) | 13 (11.2) | 3 (7.1) | 2 (11.1) | 2 (6.9) | 6 (22.2) | 0.210 |
| Previous peripheral artery disease, n (%) | 2 (1.7) | 0 (0) | 1 (5.6) | 1 (3.4) | 0 (0) | 0.351 |
| Hypercholesterolemia, n (%) | 45 (38.8) | 17 (40.5) | 5 (27.8) | 15 (51.7) | 8 (29.6) | 0.265 |
| Previous heart failure, n (%) | 5 (4.3) | 1 (2.4) | 2 (11.1) | 1 (3.4) | 1 (3.7) | 0.480 |
| Characteristics of the event |  |  |  |  |  |  |
| Duration of the event ≥ 60', n (%) | 63 (54.8) | 21 (51.2) | 8 (44.4) | 18 (62.1) | 16 (59.3) | 0.582 |
| Multiple events, n (%) | 25 (21.6) | 10 (23.8) | 4 (22.2) | 5 (17.2) | 6 (22.2) | 0.928 |
| Carotid territory event, n (%) | 70 (60.3) | 27 (64.3) | 9 (50.0) | 20 (69.0) | 14 (51.9) | 0.425 |
| Vertebrobasilar event, n (%) | 9 (7.8) | 2 (4.8) | 2 (11.1) | 1 (3.4) | 4 (14.8) | 0.328 |
| Undetermined territory event, n (%) | 37 (31.9) | 7 (38.9) | 8 (27.6) | 8 (27.6) | 9 (23.3) | 0.874 |
| Possible, non-definitive TIA event, n (%) | 4 (3.4) | 1 (2.4) | 1 (5.6) | 1 (3.4) | 1 (3.7) | 0.942 |
| Speech impairment, n (%) | 85 (73.3) | 32 (76.2) | 13 (72.2) | 21 (72.4) | 19 (70.4) | 0.956 |
| Motor impairment, n (%) | 62 (53.4) | 20 (47.6) | 12 (66.7) | 17 (58.6) | 13 (21.0) | 0.484 |
| Isolated sensory impairment, n (%) | 5 (4.3) | 3 (7.1) | 0 (0) | 1 (3.7) | 0.930 | 0.636 |
| Campimetric visual deficit, n (%) | 5 (4.3) | 1 (2.4) | 2 (11.1) | 1 (3.4) | 1 (3.7) | 0.480 |
| ABCD<sup>2</sup> score, median (IQR) | 5.0 (4.0-6.0) | 5.0 (4.0-6.0) | 5.0 (4.0-6.0) | 5.0 (4.0-6.0) | 5.0 (4.0-6.0) | 0.414 |
| Missing | 1 |  |  |  |  |  |
| CHA<sub>2</sub>DS<sub>2</sub>-VASc, mean (SD) | 3.4 (1.6) | 3.2 (1.6) | 3.8 (1.7) | 3.7 (1.3) | 3.1 (1.5) | 0.326 |
| ASCOD classification, n (%) |  |  |  |  |  |  |
| A1 or 2 | 18 (15.5) | 3 (7.1) | 4 (22.2) | 6 (20.7) | 5 (18.5) | 0.303 |
| S1 or 2 | 7 (6.0) | 0 (0) | 1 (5.6) | 2 (6.9) | 4 (14.8) | 0.093 |
| Neuroimaging features |  |  |  |  |  |  |
| Positive DWI, n (%) | 58 (59.8) | 22 (61.1) | 9 (64.3) | 12 (50.0) | 15 (65.2) | 0.710 |
| Missing | 19 |  |  |  |  |  |
| Pattern of DWI lesion, n (%) |  |  |  |  |  |  |
| Scattered in one vascular territory | 22 (37.3) | 9 (39.1) | 5 (55.6) | 4 (33.3) | 4 (26.7) | 0.548 |
| Cortical lesion in one vascular territory | 24 (40.7) | 11 (47.8) | 2 (22.2) | 6 (50.0) | 5 (33.3) | 0.471 |
| Multiple vascular territories | 5 (8.5) | 3 (13.0) | 1 (11.1) | 1 (8.3) | 0 (0) | 0.554 |
| Subcortical | 8 (13.6) | 0 (0) | 1 (11.1) | 1 (8.3) | 6 (40.0) | 0.005 |
| Follow-up events |  |  |  |  |  |  |
| Ischemic stroke recurrence, n (%) | 21 (18.1) | 4 (9.5) | 4 (22.2) | 10 (34.5) | 3 (11.1) | 0.038 |
| Biomarker results; median (IQR) |  |  |  |  |  |  |
| NSE, pg/ml | 7.5 (5.0-10.3) | 7.5 (5.0-10.2) | 7.9 (5.4-10.5) | 6.5 (3.7-9.4) | 7.1 (4.5-9.4) | 0.876 |
| Hs-CRP, mg/l | 2.7 (1.3-13.0) | 3.0 (1.3-10.9) | 7.3 (1.7-10.9) | 4.7 (1.1-9.2) | 1.7 (0.8-5.1) | 0.243 |
| IL-6, pg/ml | 4.6 (2.0-7.3) | 4.6 (2.6-6.3) | 6.8 (1.5-12.1) | 3.3 (2.8-6.4) | 3.7 (1.7-12.3) | 0.361 |
| NT-proBNP, pg/ml | 466.4 (218.7-1031.0) | 602.7 (424.6-1885.5) | 602.7 (382.8-1861.0) | 614.2 (197.4-1031.0) | 211.8 (202.3-263.6) | 0.006 |
| Neopterin, nmol/l | 11.9 (9.6-20.2) | 13.9 (9.2-19.5) | 12.0 (6.0-17.9) | 24.6 (10.0-39.2) | 11.1 (10.3-19.6) | 0.400 |
| Copeptin, pmol/l | 9.4 (5.8-10.5) | 10.5 (8.0-15.6) | 9.9 (9.3-10.5) | 8.9 (8.3-9.4) | 5.3 (3.9-7.8) | 0.150 |
| Adiponectin, microg/ml | 11.7 (8.2-17.8) | 11.5 (8.0-19.6) | 12.0 (11.7-12.3) | 12.4 (12.1-12.6) | 8.9 (8.0-23.0) | 0.858 |
| TNF-α, pg/ml | 93.0 (69.0-151.0) | 113.0 (84.5-146.5) | 71.0 (49.0-93.0) | 116.5 (59.0-174.0) | 94.5 (71.5-145.5) | 0.587 |
| IL-1 $\alpha$, pg/ml | 109.0 (75.0-169.0) | 119.0 (100.5-160.0) | 81.5 (57.0-106.0) | 130.5 (70.0-191.0) | 93.5 (69.0-152.0) | 0.577 |
| S100b, pg/ml | 31.8 (23.7-65.1) | 31.8 (27.2-57.1) | 21.4 (16.7-26.0) | 24.1 (23.7-24.6) | 52.8 (36.1-81.3) | 0.556 |

IQR, interquartile range; SD, standard deviation; DWI, diffusion-weighted imaging; ASCOD (A, atherosclerosis; S, small vessel disease; C, cardiac pathology; O, other causes; and D, dissection) classification. hs-CRP, high-sensitivity C-reactive protein; IL, interleukin; TNF-α , tumor necrosis factor-alpha; NSE, neuron-specific enolase; NT-proBNP, N-terminal pro-B type natriuretic peptide
\* Statistical significance after Bonferroni adjustment

### Predictors of new-onset atrial fibrillation

After the exclusion of previous AF patients, new-onset AF was associated with sex (female) (hazard ratio [HR] 1.61 [95% CI, 1.07– 2.41]; P=0.013), age (HR 1.05 [95% CI, 1.03–1.07]; P<0.001), previous ischemic heart disease (HR 1.84 [95% CI 1.15-2.97]; P=0.012), and cortical DWI lesion pattern (HR 2.81 [95% CI 1.87-4.21]; P<0.001) (Table 3). A cut-off value was obtained for NT-proBNP of 218.3 pg/ml, with a sensitivity of 80.6 % and a specificity of 60.5%.

**Table 3.** Cox proportional hazards regression model to assess risk of atrial fibrillation after TIA.

| Variable | Model 1 |  | Model 2 |  | Model 3 |  | Model 4 |  |
| --- | --- | --- | --- | --- | --- | --- | --- | --- |
|  | HR (95% CI) | P value | HR (95% CI) | P value | HR (95% CI) | P value | HR (95% CI) | P value |
| Sex female | 1.60 (1.10-2.31) | 0.013 | 1.61 (1.07-2.41) | 0.022 | ... | ... | ... | ... |
| Age | 1.05 (1.03-1.07) | <0.001 | 1.05 (1.03-1.08) | <0.001 | 1.07 (1.01-1.13) | 0.028 | 1.06 (1.00-1.13) | 0.042 |
| Previous IHD | 2.04 (1.32-3.16) | 0.001 | 1.84 (1.15-2.97) | 0.012 | ... | ... | ... | ... |
| Carotid territory event | 1.45 (0.98-2.14) | 0.064 | 1.88 (1.23-2.87) | 0.003 | ... | ... | ... | ... |
| S0 or 3 ASCOD classification | 2.80 (1.27-6.14) | 0.010 | ... | ... | 5.19 (0.70-38.50) | 0.107 | 5.10 (0.69-37.80) | 0.111 |
| Non-subcortical DWI pattern | ... | ... | 2.81 (1.87-4.21) | <0.001 | ... | ... | ... | ... |
| NT proBNP>218.2 pg/ml | ... | ... | ... | ... | 4.20 (1.51-11.67) | 0.006 | 4.10 (1.46-11.51) | 0.007 |

DWI, diffusion-weighted imaging; HR, hazard ratio; IHD, ischemic heart disease; ASCOD (A, atherosclerosis; S, small vessel disease; C, cardiac pathology; O, other causes; and D, dissection) classification

In the Kaplan-Meier analysis (Figure 2), female patients (log-rank test P=0.002), patients with previous ischemic heart disease (IHD) (log-rank test P=<0.001), patients with carotid territory events (log-rank test P=0.003), and patients ≥ 75 years of age (log-rank test P<0.001) had a significantly higher risk of new-onset AF throughout follow-up. In contrast, cortical DWI lesion patterns (log-rank test P<0.001) and NT-proBNP ≥218.2 pg/ml (log-rank test P<0.001) were associated with a significant risk of new-onset AF during the early follow-up, specially during the first 3-5 years.

**FIGURE 2.**
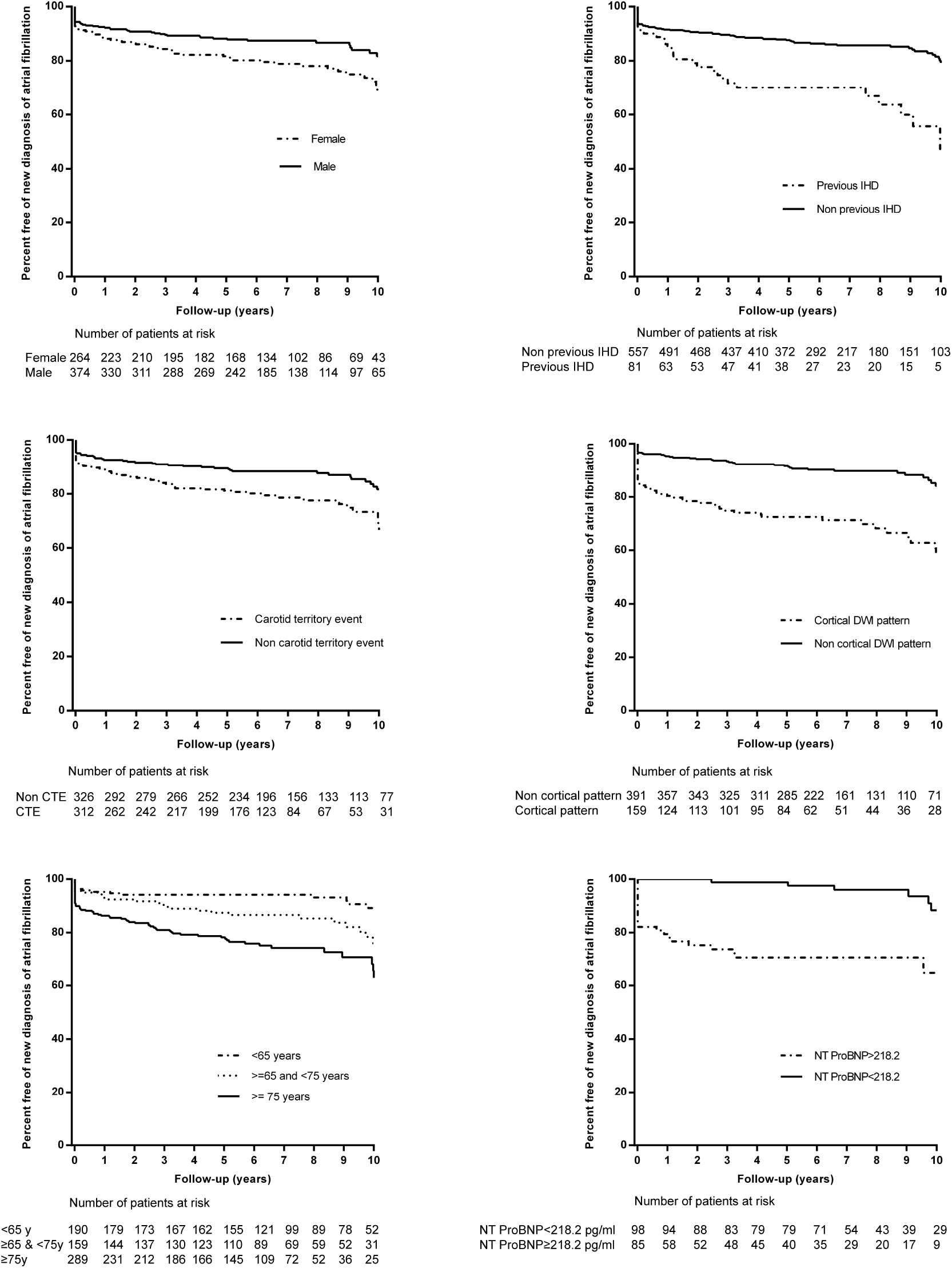
Kaplan-Meier event curves for the risk of new diagnosis of atrial fibrillation (AF) among patients with no previously diagnosed AF according to sex, previous ischemic heart disease (IHD), age, carotid territory event (CTE), diffusion-weighted imaging (DWI) pattern and NT-proBNP levels.

### New-onset AF and risk of recurrent stroke

Patients with a new diagnosis of AF after admission and before five years of follow-up had a higher risk of SR than patients with new-onset AF beyond five years of follow-up or during admission and than patients with previous AF or without new diagnosis of AF (log-rank test P=0.002) (Figure 3).

**FIGURE 3.**
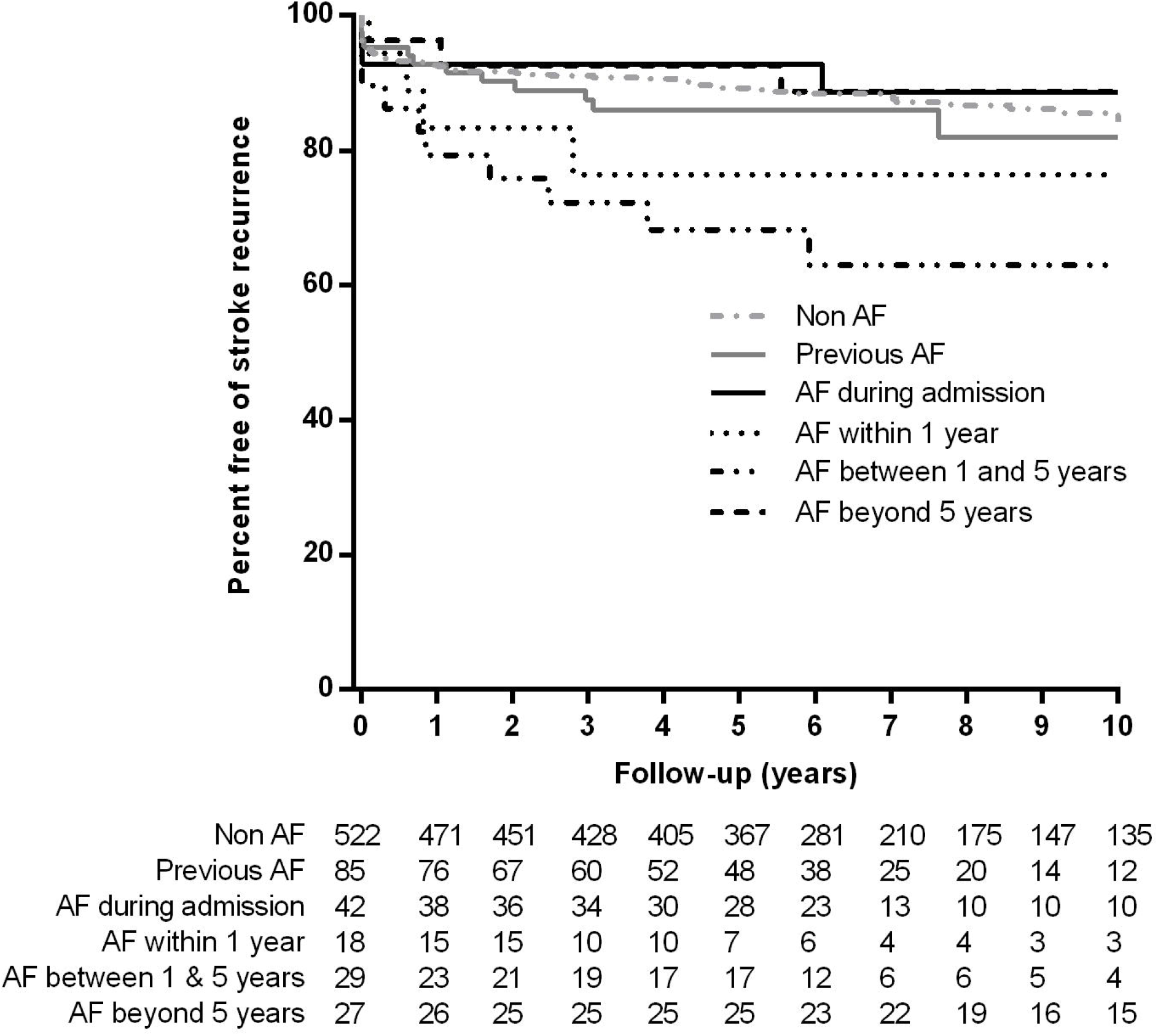
Kaplan-Meier event curves for the risk of stroke recurrence according to previous or new diagnosis of atrial fibrillation.

## DISCUSSION

In our study of high-risk patients with TIA attended at an emergency department, we observed a nonnegligible risk of new diagnosis of AF during the long follow-up. Nearly one out of five patients without previous diagnosis of AF developed a new-onset AF. Interestingly, we identified clinical, neuroimaging and biomarker predictors of new-onset AF that could be used to indicate patients who would benefit from long-term ECG monitoring. Some of these predictors differed from clinical and neuroimaging features of previous AF patients. In addition, patients with a new diagnosis of AF beyond five years of follow-up differed in the etiological phenotype of the index event and in the pattern of biomarker levels from early diagnosis AF-groups.

Our documented risk of AF was significantly higher than the risk observed in a previous meta-analysis that included studies with a limited follow-up^11^, but similar to the multicenter TIA registry.org project that reached five years of follow-up. Although AF-related brain ischemic events have been associated with disability strokes^5 26^, our results suggest the need to investigate occult AF in TIA patients with suspicious embolic events. This is especially so when we take into account that delayed diagnosis is related to an increased risk of SR as correct preventive strategies are also delayed. As in previous studies, we observed that the risk of new diagnosis of AF was related to age^26-30^, previous IHD^30^ and DWI patterns^31^. We also identified sex differences in the proportion of new-onset AF. AF was more frequent among females^32^ as the proportion of vascular risk factors is lower than in men^18^. Patients with previously described predictors and high levels of NT-ProBNP in the absence of evidence of small vessel disease would clearly benefit from exhaustive ECG monitoring. The correlation between AF and high levels of this biomarker is well known among stroke and TIA patients^25 33^. This correlation is explained by the association between pro-BNP and atrial dilatation^33^. In this regard, it should be noted that in our study patients with previous AF had higher levels of NT-proBNP than patients with new-onset AF. This should be taken into account when calculating the cut-off levels in patients with suspected cardioembolic events.

The study of cryptogenic events is a challenge. An AF episode was diagnosed during the follow-up in only one out of five patients with this condition after admission work-up. This result is in line with the failure of the RESPECT-ESUS and NAVIGATE-ESUS trials^7 8^ in demonstrating any efficacy in the prevention of SR with direct non-vitamin K agonist oral anticoagulants in patients with cryptogenic events. In these patients, arteriogenic embolism due to non-stenotic plaques or aortic arch atheroma could be the cause of the ischemic events^34^.

Our study has some relevant limitations. First, the registry was designed with SR and not the diagnosis of new AF as the main endpoint. In this sense, ECG and echocardiographic abnormalities of atrial myopathy like left atrial enlargement or P-wave abnormalities^35 36^ were not registered. Second, a larger sample size would have better guaranteed the extrapolation of our results. Third, although there were no significant differences between patients included in the biomarker substudy and patients not included, it would have been interesting to have that information for all the patients.

In conclusion, the risk of new diagnosis of AF after TIA is clinically relevant. Old age, sex-female, previous IHD, carotid territory symptoms, cortical DWI pattern lesion, absence of evidence of small vessel disease and high levels of NT-ProBNP increase the likelihood of new AF. Our results can be used to evaluate the benefit of long-term cardiac monitoring in selected patients.

