## Supplemental table 1 for "Early and long-term risk of new-onset atrial fibrillation after transient ischemic attack"

Supplementary Table I. Clinical characteristics, neuroimaging features and outcomes by new diagnosis of atrial fibrillation.

|  | **All** | **No BK determined** | **BK determined** | **p-value** |
| --- | --- | --- | --- | --- |
| Total, n (%) | 723 | 515 (71.2) | 208 (28.8) | … |
| Years of follow-up, median (IQR) | 6.5 (5.0-9.6) | 6.3 (4.8-9.3) | 7.2 (5.4-10.0) | 0.005 |
| Sex female, n (%) | 302 (41.8) | 215 (41.7) | 87 (41.8) | 0.984 |
| Age, mean (SD) years | 70.7 (11.9) | 70.9 (12.0) | 70.4 (12.0) | 0.656 |
| Previous ischemic stroke n (%) | 66 (9.1) | 38 (7.4) | 28 (13.5) | 0.010 |
| Hypertension | 481 (66.5) | 346 (67.2 | 135 (64.9) | 0.556 |
| Alcoholism, n (%) | 21 (2.9) | 16 (3.1) | 5 (2.4) | 0.610 |
| Previous ischemic heart disease, n (%) | 99 (13.7) | 75 (14.6) | 24 (11.5) | 0.284 |
| Diabetes mellitus, n (%) | 215 (29.7) | 154 (29.9) | 61 (29.3) | 0.878 |
| Active smoking, n (%) | 102 (14.1) | 73 (14.2) | 29 (13.9) | 0.935 |
| Previous atrial fibrillation, n (%) | 85 (11.8) | 63 (12.2) | 22 (10.6) | 0.531 |
| Previous peripheral artery disease, n (%) | 26 (3.6) | 21 (4.1) | 5 (2.4) | 0.274 |
| Hypercholesterolemia, n (%) | 246 (34.0) | 175 (34.0) | 71 (34.1) | 0.968 |
| Previous heart failure, n (%) | 31 (4.3) | 21 (4.1) | 10 (4.8) | 0.661 |
| Characteristics of the event |  |  |  |  |
| Systolic arterial pressure, mean (SD) mmHg | 152.8 (28.2) | 151.5 (27.9) | 155.8 (28.9) | 0.073 |
| Diastolic arterial pressure, mean (SD) mmHg | 80.0 (13.1) | 79.4 (12.7) | 81.5 (14.1) | 0.064 |
| Duration of the event, n (%) |  |  |  |  |
| ≤ 10’ | 86 (12.1) | 74 (14.6) | 12 (5.9) | 0.002* |
| 10’ to 60’ | 248 (34.9) | 164 (32.4) | 84 (41.0) |  |
| ≥ 60’ | 377 (53.0) | 268 (53.0) | 109 (53.2) |  |
| Missing | 12 |  |  |  |
| Multiple events, n (%) | 165 (22.8) | 115 (22.3) | 50 (24.0) | 0.620 |
| Carotid territory event, n (%) | 367 (50.8) | 264 (51.3) | 103 (49.5) | 0.671 |
| Vertebrobasilar event, n (%) | 78 (10.8) | 59 (11.5) | 19 (9.1) | 0.362 |
| Undetermined territory event, n (%) | 286 (39.6) | 196 (38.1) | 90 (43.3) | 0.195 |
| Possible, not definitive TIA event, n (%) | 79 (10.9) | 51 (9.9) | 28 (13.5) | 0.165 |
| Speech impairment, n (%) | 449 (62.1) | 321 (62.3) | 128 (61.5) | 0.843 |
| Motor impairment, n (%) | 377 (52.1) | 270 (52.4) | 107 (51.4) | 0.810 |
| Isolated sensory impairment, n (%) | 53 (7.3) | 36 (7.0) | 17 (8.2) | 0.581 |
| Campimetric visual deficit, n (%) | 22 (3.0) | 18 (3.5) | 4 (1.9) | 0.343 |
| ABCD2 score, median (IQR) | 5.0 (4.0-6.0) | 5.0 (4.0-6.0) | 5.0 (4.0-6.0) | 0.987 |
| Missing | 12 |  |  |  |
| CHA2DS2-VASC, mean (SD) | 3.0 (1.6) | 2.7 (1.7) | 3.7 (1.3) | 3.4 (1.6) |
| ASCOD Grades, n (%) |  |  |  |  |
| A1 or 2 | 144 (19.9) | 95 (18.4) | 49 (23.6) | 0.119 |
| A0 or 3 | 579 (80.1) | 420 (81.6) | 159 (76.4) |  |
| S1 or 2 | 109 (15.1) | 74 (14.4) | 35 (16.8) | 0.403 |
| S0 or 3 | 614 (84.9) | 441 (85.6) | 173 (83.2) | 0.403 |
| Neuroimaging features |  |  |  |  |
| Positive DWI, n (%) | 244 (39.7) | 156 (37.4) | 88 (44.7) | 0.086 |
| Follow-up events |  |  |  |  |
| Ischemic stroke recurrence, n (%) | 98 (13.6) | 67 (13.0) | 31 (14.9) | 0.501 |

IQR, interquartile range; SD, standard deviations; DWI, diffusion weighted imaging

* Statistical significance after Bonferroni adjustment

Table 2.
